# Comparison of Large Language Models in Answering Immuno-Oncology Questions: A Cross-Sectional Study

**DOI:** 10.1101/2023.10.31.23297825

**Authors:** Giovanni Maria Iannantuono, Dara Bracken-Clarke, Fatima Karzai, Hyoyoung Choo-Wosoba, James L. Gulley, Charalampos S. Floudas

**Affiliations:** Genitourinary Malignancies Branch, Center for Cancer Research, National Cancer Institute, National Institutes of Health, Bethesda, MD, United States; Center for Immuno-Oncology, Center for Cancer Research, National Cancer Institute, National Institutes of Health, Bethesda, MD, United States; Biostatistics and Data Management Section, Center for Cancer Research, National Cancer Institute, National Institutes of Health, Bethesda, MD, United States

**Author notes:** These authors contributed equally to this work (Giovanni Maria Iannantuono, Dara Bracken-Clarke). **Corresponding Author:** Charalampos S. Floudas, MD, DMSc, MS. Center for Immuno-Oncology, Center for Cancer Research, National Cancer Institute, 10 Center Drive, Bethesda, MD, 20892. Building 10, Room 7N240A, - *Email:. Author Contributions Iannantuono GM: *Conception/Design; Provision of study material*; *Collection and/or assembly of data; Data analysis and interpretation; Manuscript writing; Final approval of manuscript.* Bracken-Clarke D: *Conception/Design; Provision of study material*; *Collection and/or assembly of data; Data analysis and interpretation; Manuscript writing; Final approval of manuscript*. Karzai F: *Manuscript writing; Final approval of manuscript*. Choo-Wosoba H:*Data analysis and interpretation; Manuscript writing; Final approval of manuscript*. Gulley JL: *Manuscript writing; Final approval of manuscript*. Floudas CS: *Conception/Design; Data analysis and interpretation; Manuscript writing; Final approval of manuscript*.

**Keywords:** Large language models, Artificial intelligence, Immuno-oncology, ChatGPT, Google Bard

## Abstract

**Background:** The capability of large language models (LLMs) to understand and generate human-readable text has prompted the investigation of their potential as educational and management tools for cancer patients and healthcare providers.

**Materials and Methods:** We conducted a cross-sectional study aimed at evaluating the ability of ChatGPT-4, ChatGPT-3.5, and Google Bard to answer questions related to four domains of immuno-oncology (Mechanisms, Indications, Toxicities, and Prognosis). We generated 60 open-ended questions (15 for each section). Questions were manually submitted to LLMs, and responses were collected on June 30th, 2023. Two reviewers evaluated the answers independently.

**Results:** ChatGPT-4 and ChatGPT-3.5 answered all questions, whereas Google Bard answered only 53.3% (p <0.0001). The number of questions with reproducible answers was higher for ChatGPT-4 (95%) and ChatGPT3.5 (88.3%) than for Google Bard (50%) (p <0.0001). In terms of accuracy, the number of answers deemed fully correct were 75.4%, 58.5%, and 43.8% for ChatGPT-4, ChatGPT-3.5, and Google Bard, respectively (p = 0.03). Furthermore, the number of responses deemed highly relevant was 71.9%, 77.4%, and 43.8% for ChatGPT-4, ChatGPT-3.5, and Google Bard, respectively (p = 0.04). Regarding readability, the number of highly readable was higher for ChatGPT-4 and ChatGPT-3.5 (98.1%) and (100%) compared to Google Bard (87.5%) (p = 0.02).

**Conclusion:** ChatGPT-4 and ChatGPT-3.5 are potentially powerful tools in immuno-oncology, whereas Google Bard demonstrated relatively poorer performance. However, the risk of inaccuracy or incompleteness in the responses was evident in all three LLMs, highlighting the importance of expert-driven verification of the outputs returned by these technologies.

**IMPLICATIONS FOR PRACTICE:** Several studies have recently evaluated whether large language models may be feasible tools for providing educational and management information for cancer patients and healthcare providers. In this cross-sectional study, we assessed the ability of ChatGPT-4, ChatGPT-3.5, and Google Bard to answer questions related to immuno-oncology. ChatGPT-4 and ChatGPT-3.5 returned a higher proportion of responses, which were more accurate and comprehensive, than those returned by Google Bard, yielding highly reproducible and readable outputs. These data support ChatGPT-4 and ChatGPT-3.5 as powerful tools in providing information on immuno-oncology; however, accuracy remains a concern, with expert assessment of the output still indicated.

## 1. INTRODUCTION

Large language models (LLMs) are a recent breakthrough in the domain of generative artificial intelligence (AI) (1). Generative AI includes technologies based on “natural language processing” (NLP) which uses computational linguistics and deep learning (DL) algorithms to enable computers to interpret and generate human-like text (2). Large language models are complex systems trained on large quantities of text data which are able to create new content in response to prompts such as text, images, or other media (3). This versatility has led to the investigation of their potential applications in the field of medicine and healthcare in light of its self-evident potential benefits in these domains (4). Indeed, the availability of user-friendly tools able to provide detailed, accurate and current information would be crucial in promoting patient and healthcare providers’ education and awareness, particularly in the case of complex health conditions like cancer (5).

Thus far, many studies have assessed the potential of ChatGPT, an advanced LLM based on a generative pre-trained transformer (GPT) architecture, for providing screening and/or management information in solid tumors (6). Following the rollout of ChatGPT, more LLMs trained on different data were released, expanding the selection of these new AI-based tools. Consequently, an increasing number of studies are investigating and comparing the potential ability of ChatGPT with other LLMs as easy-to-use interfaces to gather information related to a specific cancer-related topic (7). So far, initial evidence suggests a possible role of these technologies as “virtual assistants” for healthcare professionals and patients in providing information about cancer, unfortunately counterbalanced by a significant error rate. Therefore, further studies are needed to investigate the potential applicability of these tools in other fields (7).

The past several years have seen profound changes in the field of immuno-oncology (IO). The advent of immune-checkpoint inhibitors (ICIs) has paved the way towards a new era in cancer treatment, enhancing the chance of long-term survival in patients with metastatic disease, and providing new treatment options in earlier-stage settings (8). Presently, an increasing number of cancer patients are either candidates for or already receiving ICIs or other immunotherapies, subject to both the enormous potential benefits but also the immune-related adverse events that may be caused by these treatments (9). In this context, LLMs may represent a valid tool for healthcare professionals and patients (and their caregivers) receiving these treatments. Therefore, we sought to assess and compare the ability of three prominent LLMs to provide educational and management information in the IO field.

## 2. MATERIALS AND METHODS

### 2.1 Large language models

In this cross-sectional study we compared the performance of three LLMs: ChatGPT-3.5 (10), ChatGPT-4 (10), and Google Bard (11). ChatGPT is an LLM based on the GPT architecture and developed by OpenAI, a company based in San Francisco (USA). ChatGPT is built upon either GPT-3.5 and GPT-4; the former is freely available to all the users, whereas the latter is an advanced version with additional features and provided under the name “ChatGPT Plus” to paid subscribers (10). Google Bard is based on the Pathways Language Model (PaLM) family of LLMs, developed by GoogleAI (11).

### 2.2 Questions and responses’ generation

We generated 60 open-ended questions based on our clinical experience covering four different domains of IO including “mechanisms” (of action), “indications” (for use), “toxicities”, and “prognosis” (Suppl. Mat. A). In order to standardize assessment, particularly of “relevance” and “accuracy”, and to reduce bias, a sample answer for each question was generated *a priori* prior to question submission. Questions were manually and directly submitted to the web chat interfaces of the three above-mentioned LLMs on June 30th 2023 and responses were collected (Suppl. Mat. B). We assessed the reproducibility, accuracy, relevance, and readability (Table 1) of responses provided by each LLM. Two reviewers (GMI and DBC) rated the answers independently. During the rating process, reviewers were blinded to the LLM being assessed. Inconsistencies between the reviewers were discussed with an additional reviewer (CSF) and resolved by consensus. Cohen’s kappa coefficient was calculated to evaluate inter-rater reliability during the rating process (12).

**Table 1.** Definitions of the outcomes.

| **Outcomes** | **Definitions** | **Score** |
| --- | --- | --- |
| **Answer Returned** | *The ability of LLM to return a meaningful answer to each instance of the question submitted, rather than returning an error or declining to return an answer, independent of the accuracy of this response.* | Recorded as Boolean True/False |
| **Reproducibility** | *The ability of LLM to return a generally similar series of answers across the three separate queries with no fundamental differences or inconsistencies between these three answers.* | Recorded as Boolean True/False |
| **Accuracy** | *The ability of LLM to provide accurate and correct information addressing the question asked and returning all major or critical points required in such an answer. Response <u>not</u> adversely marked for extraneous or irrelevant information here – as long as this information was correct.* | Recorded numerically from 1 to 3 |
| **Readability** | *The ability of LLM to return comprehensible and coherent natural language text in English, including appropriate syntax, formatting, and punctuation, independent of the accuracy of this response.* | Recorded numerically from 1 to 3 |
| **Relevance** | *The ability of LLM to return information that was relevant and specific to the question asked or immediately adjacent topics without extraneous, unrequested, or tangential information. Accuracy was not specifically assessed here, though the result was adversely marked if the response included immaterial information while neglecting to address the specific question asked.* | Recorded numerically from 1 to 3 |

Note: for scoring of Relevance, the answer returned was not adversely marked for any included disclaimers to the effect that the LLM cannot provide medical advice and any such advice should be sought from a clinician or that anyone with a cancer diagnosis and/or receiving systemic therapy with potential toxicity should contact their treating clinician/s. This was deemed to represent appropriate and medically sound advice and not to be irrelevant or extraneous material.

First, we assessed the ability of each LLM to provide reproducible responses. Therefore, each individual question was submitted three times on each LLM. In the case of non-reproducible answers, questions were not considered for further analysis. Subsequently, the accuracy, relevance, and readability of responses deemed reproducible were assessed using a 3-point scale (Table 2) (Figure 1). Reviewers graded the accuracy of answers according to available information as of 2021, as the training datasets of ChatGPT are updated to September 2021. Finally, word- and character-counts were calculated for each answer.

**Table 2.** Definitions of the scoring system.

|  | **Score** |  |  |
| --- | --- | --- | --- |
|  | **1** | **2** | **3** |
| **Accuracy** | *Fundamentally inaccurate or incorrect information, including critical errors, omissions and/or entirely incorrect treatment advice.* | *Partially correct and accurate information, including non-critical errors and/or omitting relevant information or failing to provide specific guideline advice.* | *Fully accurate and correct information, answering the specific question asked with no significant errors or omissions.* |
| **Relevance\*** | *Irrelevant and/or entirely tangential material, not addressing the specific question asked.* | *Generally relevant material though including significant extraneous and/or tangential information.* | *Relevant and focused information directly addressing the question asked, including an appropriate expansion on the relevant topic.* |
| **Readability** | *Incoherent, unintelligible and/or garbled text, +/- severely misformatted and/or oxymoronic material resulting in compromised legibility.* | *Generally coherent and intelligible material with significant formatting and/or parsing errors.* | *Fully coherent, well-parsed and constructed material, easily and clearly intelligible.* |

\*Note: Inclusion of a disclaimer that the answer was provided by an AI/LLM and cannot be taken as medical advice and/or that any information or questions should also be addressed to a qualified medical practitioner was not scored negatively – as this represents a legitimate and appropriate legal disclaimer.

**Figure 1:**
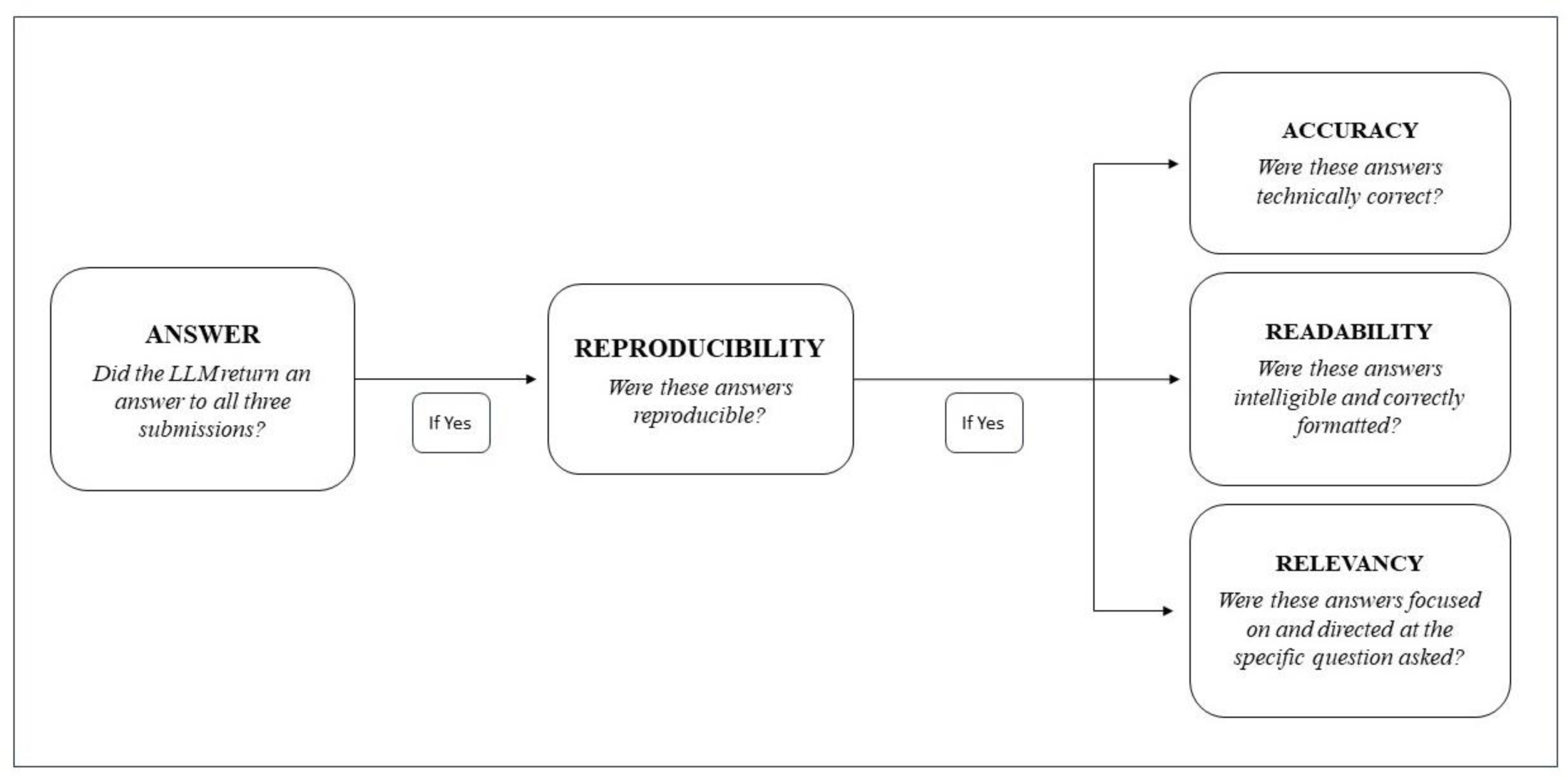
Flowchart of the rating process for each triplet of responses.

### 2.3 Statistical analyses

Categorical variables were presented with proportions and numeric variables as measures of central tendency. Comparisons between categorical variables were performed with two-sided generalized Fisher’s exact tests for testing any potential differences in these three LLMs. In the case of numeric continuous variables, a Kruskal-Wallis test was utilized. Statistical tests were not performed within each of the four domains, but rather were performed only to evaluate overall performance by combining those four domains, due to insufficient sample sizes within each domain (i.e., only up to 15 available observations). All statistical results should be interpreted as exploratory; all statistical analyses were performed and all plots generated using R version 4.2.2 (The R Foundation for Statistical Computing, 2022). This study was conducted in accordance with Strengthening the Reporting of Observational Studies in Epidemiology (STROBE) reporting guidelines (13).

## 3. RESULTS

Assessment of inter-rater reliability with Cohen’s kappa during the rating process demonstrated “strong” to “near perfect” agreement between reviewers (Suppl. Mat. C). ChatGPT-3.5 and ChatGPT-4 provided at least one response to all questions (60 [100%]), while Google Bard responded only to 32 (53.3%) queries (p <0.0001). Specifically, the percentages of responses provided by Google Bard were different across the four domains, with better performances in the “mechanisms” (14 [93.3%]) and “prognosis” domains (13 [86.7%]) compared to the “indications” (5 [33.3%]), and “toxicities” (0 [0%]) domains. Regarding reproducibility, the numbers of questions with reproducible answers were similar between ChatGPT-3.5 and ChatGPT-4 (53 [88.3%] and 57 [95%], respectively), while it was lower (16 [50%]) for Google Bard (p <0.0001). Although ChatGPT-3.5 and ChatGPT-4 performed similarly across all domains, ChatGPT-4 achieved 100% reproducible responses in two domains (“mechanisms” and “indications”) in which ChatGPT-3.5 achieved only 86.7%. Google Bard was variably capable and accurate across the different sections. Despite a significant number of answers deemed reproducible in the “mechanisms” (6 [40%]) and “prognosis” (9 [60%]) sections, a poor performance was observed in the “indications” (1 [6.7%]) and “toxicities” (0 [0%]) domains (Figure 2). In terms of accuracy, the numbers of answers deemed fully correct were 31 (58.5%), 43 (75.4%), and 7 (43.8%) for ChatGPT-3.5, ChatGPT-4 and Google Bard, respectively (p = 0.03). Furthermore, regarding relevancy, the numbers of responses deemed highly relevant were 41 (77.4%), 41 (71.9%), and 7 (43.8%) for ChatGPT-3.5, ChatGPT-4 and Google Bard, respectively (p = 0.04). Readability was deemed optimal across all three LLMs. However, the numbers of highly readable answers were greater for ChatGPT-3.5 and ChatGPT-4 (52 [98.1%] and 57 [100%]) compared to Google Bard (14 [87.5%]) (p = 0.02) (Figure 3). The median numbers of words and their corresponding ranges for the responses provided by ChatGPT-3.5, ChatGPT-4, and Google Bard were 297 (197 - 404), 276 (139 - 395), and 290.5 (12 - 424), respectively (p = 0.06). Finally, the median numbers of characters and their corresponding ranges were 1829 (1119 - 2470), 1589 (854 - 2233), and 1532 (75 - 2070), respectively (p <0.0001).

**Figure 2:**
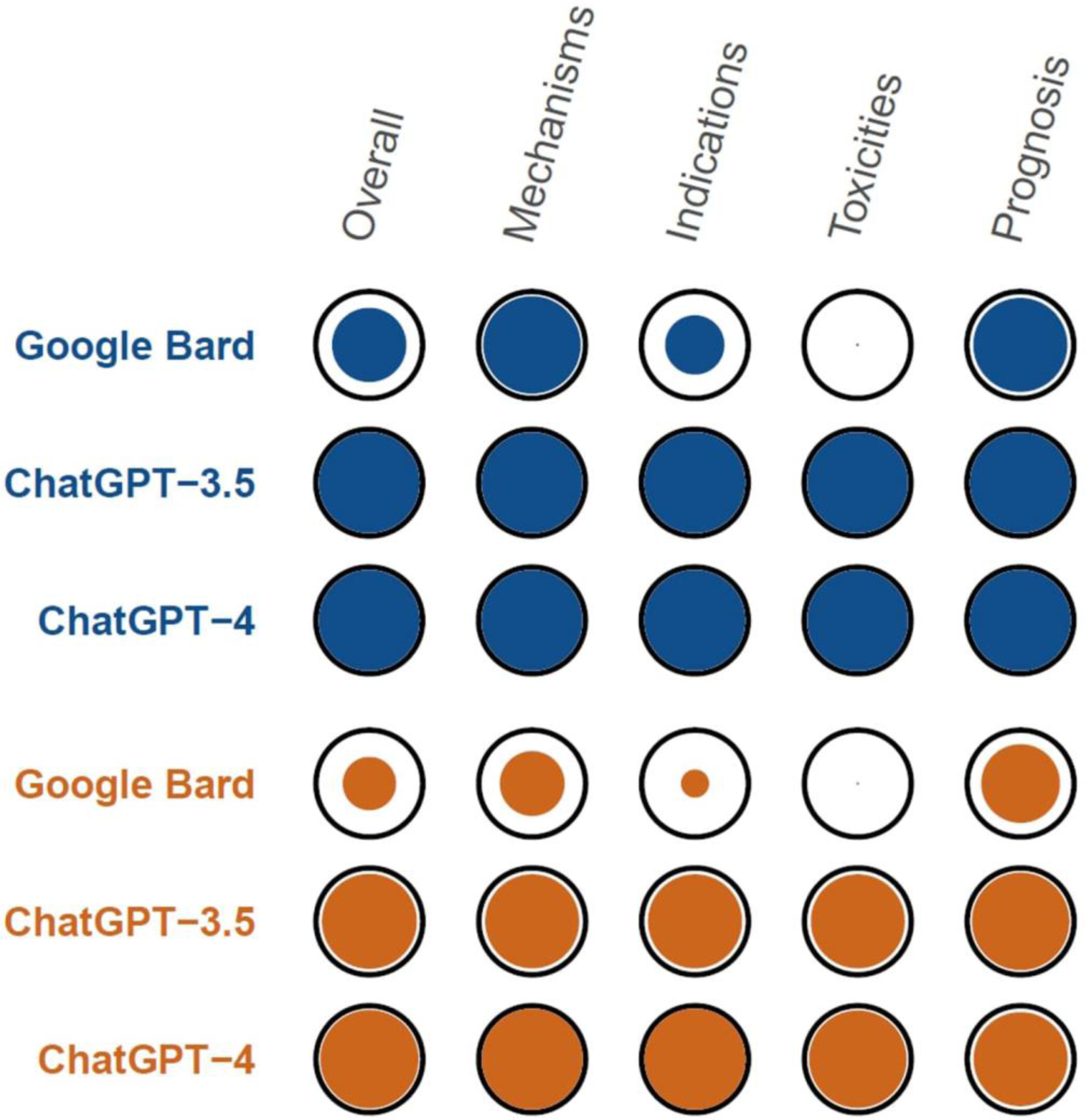
Spot matrix of the percentages of the answered questions [Blue] and reproducible responses [Orange] for each LLM. Color volume is directly proportional to percentage with the outer black circle representing 100%. Corresponding numeric data are available in Suppl. Mat. D.

**Figure 3:**
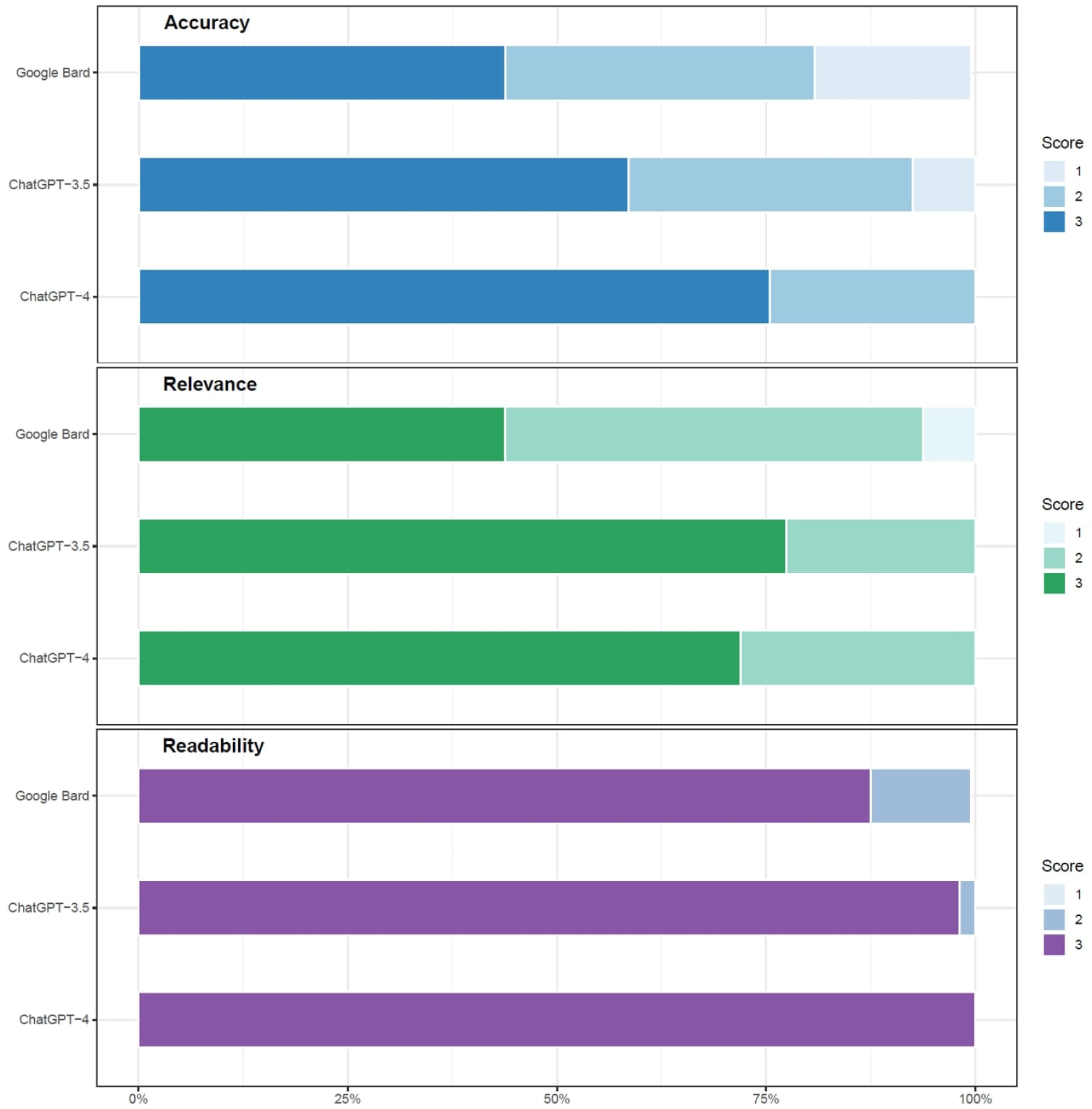
Bar plot of the results (accuracy, readability, and relevance) for all three LLMs. This plot was based only on the questions evaluable for accuracy, readability, and relevance. Corresponding numeric data are available in Suppl. Mat. D.

## 4. DISCUSSION

In recent decades, significant effort has been made to harness the potential of AI in medicine and healthcare (14). Artificial intelligence can be defined as “the science and engineering of making intelligent machines, especially intelligent computer programs” (15). It is composed of multiple subfields, based on different algorithms and principles, including knowledge representation, machine learning (ML), DL, and NLP (2,16). Specifically, NLP uses computational language and DL to enable computers to understand text in the same way as humans (2). Recent progress in NLP has led to major breakthroughs in the field of generative AI, as evidenced by the advent of LLMs (3). These can recognize, summarize and generate novel content using statistical connections between letters and words. Indeed, LLMs can also be considered as “few shot learners” due to their ability to readily adapt to new domains with few information after being trained (17).

Over the last year, the release of ChatGPT (10) has attracted considerable attention, which only increased following the release of other LLMs such as Google Bard (11), Bing AI (18) and, Perplexity (19). The remarkable adaptability of these AI-based technologies to a broad and extensive range of disciplines was immediately apparent following their introduction (20). This is also evidenced by the rapid publication of large numbers of studies designed to investigate their role in multiple and diffuse fields, including medicine and healthcare. Initial data have demonstrated LLMs to be highly applicable to the field of cancer care, especially in providing information about the screening and/or management of specific solid tumors (7). However, to the authors’ knowledge, their potential role in the field of IO has not yet been investigated, despite the rapidly expanding knowledge in all the aspects of IO (basic, translational, and clinical research) and the large number of cancer patients currently receiving immunotherapy (8,9).

Therefore, we performed a cross-sectional study aimed for the first time at assessing the potential of three prominent LLMs in answering questions about the field of IO. Our results demonstrated that ChatGPT-4 and ChatGPT-3.5 were able to answer most of the IO-related questions with excellent accuracy and relevance. In contrast, the performance of Google Bard was comparatively poorer, as shown by a lower number of both answered questions and the reproducibility/accuracy of these responses, compared to the other two LLMs. All three LLMs were able to provide highly readable responses, highlighting the power of these generative AI technologies in providing human-readable text. ChatGPT (both v3.5 and v4) clearly demonstrated their potential as a “virtual assistant” for both clinicians and patients or caregivers. ChatGPT (both v3.5 and, especially, v4) has also demonstrated remarkable acumen in both diagnosing and providing management plans for IO toxicities. It has also proved highly effective in suggesting evidence-based and licensed indications for IO therapy, either alone or in combination. Additionally, it has demonstrable efficacy in providing background information on IO drug mechanisms and disease prognoses in generally comprehensible text without excess jargon, albeit often with a lack of sources and broken or inaccurate references.

However, the results of this study also highlight the differing performance of various LLMs across topics and specific tasks (Table 3), as this demonstrates significant variability. In our study, ChatGPT is demonstrated to be a powerful tool when applied to the field of IO, particularly in comparison to Google Bard. Similar results were also reported in another recently published study assessing these three LLMs in a different cancer-related topic. Specifically, Rahsepar et al. reported the results of a study investigating the ability of ChatGPT-3.5, ChatGPT-4 and Google Bard in answering questions related to lung cancer screening and prevention (21). As in our study, ChatGPT achieved a superior performance to Google Bard. However, the available evidence suggests that the LLM developed by OpenAI is not always accurate, as shown by the results of other studies investigating medical/healthcare topics other than cancer (Table 3). In the studies published by Seth et al., Zúñiga Salazar et al. and Dhanvijay et al., Google Bard performed better in comparison to ChatGPT in non-cancer domains, likely clarifying a potential role for this LLM (22–24). Furthermore, the results of the study by Al-Ashwal et al. showed a better performance for Bing AI in answering questions related to drug-drug interactions in comparison to the other LLMs (25). Therefore, it is essential to compare the performance of different LLMs since their abilities may vary based on both task and domain.

**Table 3.** List of studies investigating the utility of ChatGPT and Google Bard across various contexts of medicine and healthcare.

| **First Author** | **Year of Publication** | **LLMs** | **Domain** | **Questions (n)** | **Reviewers (n)** |
| --- | --- | --- | --- | --- | --- |
| Al-Ashwal FY (25) | 2023 | ChatGPT - Google Bard - Bing AI | Drug-drug interactions | 225 [OE] | NA |
| Dhanvijay AK (24) | 2023 | ChatGPT - Google Bard - Bing AI | Physiology | 77 [OE] | 2 |
| Seth I (22) | 2023 | ChatGPT - Google Bard - Bing AI | Rhinoplasty | 6 [OE] | 3 |
| Koga S (29) | 2023 | ChatGPT - Google Bard | Neurodegenerative disorder | 25 [OE] | NA |
| Kumari A (30) | 2023 | ChatGPT - Google Bard | Hematology | 50 [OE] | 3 |
| Lim ZW (31) | 2023 | ChatGPT - Google Bard | Myopia | 31 [OE] | 3 |
| Meo SA (32) | 2023 | ChatGPT - Google Bard | Endocrinology, diabetes, and diabetes technology | 100 [MC] | - |
| Toyama Y (33) | 2023 | ChatGPT - Google Bard | Radiology | 103 [MC] | 3 |
| Waisberg E (34) | 2023 | ChatGPT - Google Bard | Ophthalmology | NA | 4 |
| Zuniga Salazar G (23) | 2023 | ChatGPT - Google Bard - Bing AI | Emergency | 176 [OE] | NA |

Abbreviations: Multiple choice (MC); Not available (NA); Open-ended (OE).

In addition, despite the promising results of our study and its unequivocal efficacy in synthesizing and evaluating textual data, the potential of ChatGPT for error and hallucination remains (26). The occurrence of “hallucinations” is one of the greatest obstacles to the routine clinical application of LLMs. While potentially tolerable in other domains, this is a critical issue in medicine and the biomedical sciences due to its potential to directly impact patient care. In addition, it must be noted that the datasets on which these models were trained were: (i) confidential and proprietary (thus impossible to assess for data quality or bias), (ii) not specifically selected *ab initio* for addressing biomedical issues and (iii) only valid up to September 2021 (thus lacking up to date information – a major issue in so rapidly evolving a field as medicine in general and IO in particular) (10,27). Therefore, expert assessment of LLMs’ output remains a prerequisite for their clinical use.

Open-source LLMs trained on specific biomedical datasets in order to accomplish pre-specified tasks offer a potential solution and alternative paradigm. BioGPT, a cutting-edge LLM with a user-friendly interface developed for the biomedical field, represents an excellent example of this (28). BioGPT shares the same architecture as OpenAI’s GPT models but was trained on information derived from the biomedical literature. It has demonstrated excellent performance in several tasks, including text generation and categorization, due to its extensive pre-training on massive biomedical datasets (28). Further studies to investigate the utility and performance of LLMs developed on biomedical data, with comparison to those LLMs presently available, are, thus, required.

### 4.1 Limitations

Our study has some limitations that need to be mentioned. Firstly, we have focused only on three prominent LLMs, excluding other LLMs including BingAI and Perplexity. At the time of the design of this study, ChatGPT and Google Bard were the most investigated LLMs and, thus, we elected to focus on them. However, recent evidence has shown the potential of BingAI in the biomedical field. Therefore, our results do not represent the entire spectrum of LLMs available and further assessment of other LLMs in the field of IO is essential. Secondly, the rating process of the answers was made by only two reviewers. However, while a third reviewer was available to resolve any conflicts which arose, this proved unnecessary as a strong to near perfect agreement was demonstrated between the two reviewers Finally, the number of open-ended questions included was relatively small, which may have impacted the analysis, particularly for domain-specific performance.

## 5. CONCLUSION

ChatGPT-3.5 and ChatGPT-4 have demonstrated significant and clinically meaningful utility as decision- and research-aids in various subfields of IO, while Google Bard demonstrated significant limitations, especially in comparison to ChatGPT. However, the risk of inaccurate or incomplete responses was evident in all LLMs, highlighting the importance of an expert-driven verification of the information provided by these technologies. Finally, despite their potential to positively impact the field of medicine and healthcare, this study reinforced the significance of a human evaluation of LLMs in order to create reliable tools for clinical use.

## Supporting information

List of questions submitted to LLMs

List of responses provided by LLMs

Cohen's Kappa Coefficient for Inter-Rater Reliability

Numeric data reported in Figure 2 and Figure 3

## CRediT Roles

Conceptualization (GMI, DBC, CSF); Formal analysis (GMI and HCW); Investigation (GMI and DBC); Methodology (GMI and DBC); Visualization (GMI and CSF); Writing - Original Draft (GMI, DBC, HCW); Writing - Review & Editing (FK, JG, CSF); Supervision (CSF). All authors accepted the final draft of the manuscript.

## Data Availability Statement

The data underlying this article are available in the article and in its online supplementary material.

## REFERENCES

1. IBM. What is generative AI? [Internet]. 2021 [cited 2023 Oct 13]. Available from: https://research.ibm.com/blog/what-is-generative-AI

2. IBM. What is Natural Language Processing? | IBM [Internet]. [cited 2023 Oct 15]. Available from: https://www.ibm.com/topics/natural-language-processing

3. Birhane A, Kasirzadeh A, Leslie D, Wachter S. Science in the age of large language models. Nat Rev Phys [Internet]. 2023 [cited 2023 Oct 13];5(5). Available from: https://ora.ox.ac.uk/objects/uuid:9eac0305-0a9a-4e44-95f2-c67ee9eae15c

4. Ayers JW, Poliak A, Dredze M, Leas EC, Zhu Z, Kelley JB, et al. Comparing Physician and Artificial Intelligence Chatbot Responses to Patient Questions Posted to a Public Social Media Forum. JAMA Intern Med. 2023 Jun 1;183(6):589–96.

5. Risk A, Petersen C. Health information on the internet: quality issues and international initiatives. JAMA. 2002 May 22;287(20):2713–5.

6. Sallam M. ChatGPT Utility in Healthcare Education, Research, and Practice: Systematic Review on the Promising Perspectives and Valid Concerns. Healthc Basel Switz. 2023 Mar 19;11(6):887.

7. Iannantuono GM, Bracken-Clarke D, Floudas CS, Roselli M, Gulley JL, Karzai F. Applications of large language models in cancer care: current evidence and future perspectives. Front Oncol. 2023 Sep 4;13:1268915.

8. Johnson DB, Nebhan CA, Moslehi JJ, Balko JM. Immune-checkpoint inhibitors: long-term implications of toxicity. Nat Rev Clin Oncol. 2022 Apr;19(4):254–67.

9. Darvin P, Toor SM, Sasidharan Nair V, Elkord E. Immune checkpoint inhibitors: recent progress and potential biomarkers. Exp Mol Med. 2018 Dec 13;50(12):1–11.

10. OpenAI. What is ChatGPT? [Internet]. [cited 2023 Oct 13]. Available from: https://help.openai.com/en/articles/6783457-what-is-chatgpt

11. Google. Try Bard, an AI experiment by Google [Internet]. [cited 2023 Oct 13]. Available from: https://bard.google.com

12. McHugh ML. Interrater reliability: the kappa statistic. Biochem Medica. 2012;22(3):276–82.

13. von Elm E, Altman DG, Egger M, Pocock SJ, Gøtzsche PC, Vandenbroucke JP, et al. The Strengthening the Reporting of Observational Studies in Epidemiology (STROBE) statement: guidelines for reporting observational studies. J Clin Epidemiol. 2008 Apr;61(4):344–9.

14. Haug CJ, Drazen JM. Artificial Intelligence and Machine Learning in Clinical Medicine, 2023. N Engl J Med. 2023 Mar 30;388(13):1201–8.

15. McCarthy J. What Is Artificial Intelligence?

16. IBM. AI vs. Machine Learning vs. Deep Learning vs. Neural Networks: What’s the difference? [Internet]. 2023 [cited 2023 Oct 16]. Available from: https://www.ibm.com/blog/ai-vs-machine-learning-vs-deep-learning-vs-neural-networks/

17. Brown TB, Mann B, Ryder N, Subbiah M, Kaplan J, Dhariwal P, et al. Language Models are Few-Shot Learners [Internet]. arXiv; 2020 [cited 2023 Oct 16]. Available from: http://arxiv.org/abs/2005.14165

18. Microsoft. Bing AI [Internet]. [cited 2023 Oct 17]. Available from: https://www.bing.com:9943/search?showconv=1&q=bingAI&sf=codex3p&form=MA13FV

19. Perplexity AI. Perplexity [Internet]. [cited 2023 Oct 17]. Available from: https://www.perplexity.ai/

20. Thorp HH. ChatGPT is fun, but not an author. Science. 2023 Jan 27;379(6630):313.

21. Rahsepar AA, Tavakoli N, Kim GHJ, Hassani C, Abtin F, Bedayat A. How AI Responds to Common Lung Cancer Questions: ChatGPT vs Google Bard. Radiology. 2023 Jun;307(5):e230922.

22. Seth I, Lim B, Xie Y, Cevik J, Rozen WM, Ross RJ, et al. Comparing the Efficacy of Large Language Models ChatGPT, BARD, and Bing AI in Providing Information on Rhinoplasty: An Observational Study. Aesthetic Surg J Open Forum. 2023;5:ojad084.

23. Zúñiga Salazar G, Zúñiga D, Vindel CL, Yoong AM, Hincapie S, Zúñiga AB, et al. Efficacy of AI Chats to Determine an Emergency: A Comparison Between OpenAI’s ChatGPT, Google Bard, and Microsoft Bing AI Chat. Cureus. 2023 Sep;15(9):e45473.

24. Dhanvijay AKD, Pinjar MJ, Dhokane N, Sorte SR, Kumari A, Mondal H. Performance of Large Language Models (ChatGPT, Bing Search, and Google Bard) in Solving Case Vignettes in Physiology. Cureus. 2023 Aug;15(8):e42972.

25. Al-Ashwal FY, Zawiah M, Gharaibeh L, Abu-Farha R, Bitar AN. Evaluating the Sensitivity, Specificity, and Accuracy of ChatGPT-3.5, ChatGPT-4, Bing AI, and Bard Against Conventional Drug-Drug Interactions Clinical Tools. Drug Healthc Patient Saf. 2023;15:137–47.

26. Singhal K, Azizi S, Tu T, Mahdavi SS, Wei J, Chung HW, et al. Large language models encode clinical knowledge. Nature. 2023 Aug;620(7972):172–80.

27. van Dis EAM, Bollen J, Zuidema W, van Rooij R, Bockting CL. ChatGPT: five priorities for research. Nature. 2023 Feb;614(7947):224–6.

28. Luo R, Sun L, Xia Y, Qin T, Zhang S, Poon H, et al. BioGPT: generative pre-trained transformer for biomedical text generation and mining. Brief Bioinform. 2022 Nov 19;23(6):bbac409.

29. Koga S, Martin NB, Dickson DW. Evaluating the performance of large language models: ChatGPT and Google Bard in generating differential diagnoses in clinicopathological conferences of neurodegenerative disorders. Brain Pathol Zurich Switz. 2023 Aug 8;e13207.

30. Kumari A, Kumari A, Singh A, Singh SK, Juhi A, Dhanvijay AKD, et al. Large Language Models in Hematology Case Solving: A Comparative Study of ChatGPT-3.5, Google Bard, and Microsoft Bing. Cureus. 2023 Aug;15(8):e43861.

31. Lim ZW, Pushpanathan K, Yew SME, Lai Y, Sun CH, Lam JSH, et al. Benchmarking large language models’ performances for myopia care: a comparative analysis of ChatGPT-3.5, ChatGPT-4.0, and Google Bard. EBioMedicine. 2023 Sep;95:104770.

32. Meo SA, Al-Khlaiwi T, AbuKhalaf AA, Meo AS, Klonoff DC. The Scientific Knowledge of Bard and ChatGPT in Endocrinology, Diabetes, and Diabetes Technology: Multiple-Choice Questions Examination-Based Performance. J Diabetes Sci Technol. 2023 Oct 5;19322968231203987.

33. Toyama Y, Harigai A, Abe M, Nagano M, Kawabata M, Seki Y, et al. Performance evaluation of ChatGPT, GPT-4, and Bard on the official board examination of the Japan Radiology Society. Jpn J Radiol. 2023 Oct 4;

34. Waisberg E, Ong J, Masalkhi M, Zaman N, Sarker P, Lee AG, et al. Google’s AI chatbot “Bard”: a side-by-side comparison with ChatGPT and its utilization in ophthalmology. Eye Lond Engl. 2023 Sep 28;

