## Supplementary material for "Comparison of Large Language Models in Answering Immuno-Oncology Questions: A Cross-Sectional Study": List of questions submitted to LLMs

### Supplementary Material A

- List of questions submitted to ChatGPT-3.5, ChatGPT-4, and Google Bard.

| Mechanisms | |
|---|---|
| **Q1** | *Do immunotherapy drugs directly kill cancer cells?* |
| **Q2** | *What is an immune checkpoint?* |
| **Q3** | *What are nivolumab and pembrolizumab?* |
| **Q4** | *What is relatlimab?* |
| **Q5** | *What is the target of gardasil?* |
| **Q6** | *Does cancer damage the immune system?* |
| **Q7** | *What is the difference between anti-PD-1 and anti-PD-L1 treatment?* |
| **Q8** | *How does the immune system recognize cancer?* |
| **Q9** | *What is the difference between tremelimumab and durvalumab?* |
| **Q10** | *What is tumor infiltrating lymphocyte (TIL) therapy?* |
| **Q11** | *Can the immune system reach all parts of the body?* |
| **Q12** | *Why do we combine immunotherapy with chemotherapy or radiotherapy?* |
| **Q13** | *How quickly does immunotherapy work to control cancer?* |
| **Q14** | *Does combination immunotherapy with ipilimumab and nivolumab work better than single-agent immunotherapy for kidney cancer?* |
| **Q15** | *What are the mechanisms for resistance to chimeric antigen receptor (CAR-T) therapy?* |

| Indications | |
|---|---|
| **Q1** | *Does immunotherapy work for asymptomatic melanoma brain metastases?* |
| **Q2** | *Does immunotherapy work in Hodgkin lymphoma?* |
| **Q3** | *What is the first line combination treatment for metastatic head and neck cancer?* |
| **Q4** | *What is the systemic treatment for cutaneous squamous-cell carcinoma which cannot be treated with surgery or radiotherapy?* |
| **Q5** | *What is the first line treatment for metastatic PD-L1 positive non-small cell lung cancer without an actionable mutation?* |
| **Q6** | *Is immunotherapy more effective when given in the pre-surgical (neoadjuvant) setting for melanoma?* |
| **Q7** | *Does immunotherapy work for EGFR (epidermal growth factor receptor) mutated or ALK (anaplastic lymphoma kinase) fusion non-small cell lung cancer?* |
| **Q8** | *I have Lynch syndrome and prostate cancer, is there a role for immunotherapy?* |
| **Q9** | *Does pembrolizumab work for multiple myeloma?* |
| **Q10** | *What are the systemic treatment options for metastatic uveal melanoma?* |
| **Q11** | *Does immunotherapy work after surgery for colon or rectal cancers?* |
| **Q12** | *Is there a role for immunotherapy in primary brain tumors such as glioblastoma multiforme?* |
| **Q13** | *I have good prognosis kidney cancer with an IMDC (international metastatic renal cell carcinoma database consortium) risk score of zero, should I receive immunotherapy?* |
| **Q14** | *I have ovarian cancer with a POLE (DNA polymerase epsilon) mutation, is there a role for immunotherapy?* |
| **Q15** | *Is there a role for ipilimumab in metastatic melanoma patients who have previously received nivolumab?* |

| Toxicities | |
|---|---|
| **Q1** | *Is there any risk to immunotherapy treatment in a patient with latent tuberculosis?* |
| **Q2** | *Is immunotherapy safe in patients with well-controlled HIV who are receiving antiretroviral therapy?* |
| **Q3** | *A patient receiving Nivolumab develops weight loss, tremulousness, anxiety, diarrhea, and reports feeling warm all the time. Clinically she has a fine tremor, and her eyes are proptosed. What is the likely diagnosis?* |
| **Q4** | *What is the first line treatment for pembrolizumab-induced colitis presenting with eight bowel motions per day, dehydration, and fecal incontinence?* |
| **Q5** | *A patient receiving ipilimumab and nivolumab for metastatic melanoma reports progressive, patchy whitening of his skin without any pruritis or ulceration. What is the likely diagnosis?* |
| **Q6** | *A patient receiving dostarlimab for rectal cancer is reporting progressive pruritis and grittiness to his eyes along with a dry mouth since starting treatment. What is the likely cause?* |
| **Q7** | *I have well-controlled Crohn's disease treated with mesalamine only and have been diagnosed with metastatic melanoma. Can I receive immunotherapy?* |
| **Q8** | *A patient receiving atezolizumab reports a three-week history of progressive fatigue, presyncope and weight loss and has orthostatic hypotension, hyponatremia, and borderline hypoglycemia. What is the likely diagnosis?* |
| **Q9** | *A patient receiving ipilimumab reports rapid onset of weight loss, severe thirst, frequent urination and hunger, her urine dipstick demonstrates glycosuria. What is the likely diagnosis?* |
| **Q10** | *A patient with biopsy-proven grade three colitis due to nivolumab has completed three days of methylprednisolone without significant improvement. What is the next line of therapy?* |
| **Q11** | *A patient receiving nivolumab and cabozantinib for kidney cancer has developed persistent diarrhea with approximately 4 watery bowel motions per day with no incontinence. How should this be managed?* |
| **Q12** | *A patient receiving pembrolizumab for tonsillar cancer, which was previously treated with chemoradiotherapy, develops pain, erythema and mucositis with difficulty swallowing in the previous radiation port. What is the likely pathology?* |
| **Q13** | *A patient receiving pembrolizumab reports an insidious onset of progressive muscle weakness, fatigue, myalgias, leg swelling and fatigue. Laboratory tests demonstrate elevated troponin and creatine kinase, spirometry demonstrates a restrictive pattern. What is the likely diagnosis?* |
| **Q14** | *A patient receiving ipilimumab and nivolumab for metastatic melanoma reports acute onset of headache with meningism, photophobia and nausea. Clinically she has bitemporal visual field loss and orthostatic hypotension with low thyroid stimulating hormone. What is the likely diagnosis and appropriate management?* |
| **Q15** | *A patient receiving durvalumab for lung cancer develops acute new chest pain and presyncope, his electrocardiogram demonstrates new tachycardia with depression of the ST segment. What is the likely diagnosis?* |

| Prognosis | |
|---|---|
| **Q1** | *A patient with metastatic BRAF V600E mutated melanoma who is not acutely unwell or in visceral crisis presents to start systemic therapy. Is combination immunotherapy with ipilimumab and nivolumab or targeted therapy with dabrafenib or trametinib first line associated with a better prognosis?* |
| **Q2** | *A patient presents with metastatic microsatellite unstable colon cancer. What is her prognosis with immunotherapy versus chemotherapy?* |
| **Q3** | *How does PD-L1 status impact prognosis in non-small cell lung cancer?* |
| **Q4** | *How does tumor mutational burden impact prognosis in patients receiving immunotherapy?* |
| **Q5** | *What is the response rate of B-cell acute lymphoblastic leukemia to tisagenleucel?* |
| **Q6** | *A patient receiving pembrolizumab for kidney cancer develops hypothyroidism and mild colitis but continues on immunotherapy. Does this impact his prognosis?* |
| **Q7** | *How does BRCA1/2 mutation impact response to and, thus, prognosis in patients receiving immunotherapy?* |
| **Q8** | *A patient with a history of metastatic melanoma which responded to immunotherapy relapses three years post completion of her treatment. What is the likelihood of her responding to further immunotherapy?* |
| **Q9** | *What is the impact of sipuleucel-T therapy on prostate cancer prognosis?* |
| **Q10** | *A patient with PD-L1 positive squamous cell lung cancer experiences a complete radiological response to pembrolizumab and completes two years total of therapy. What is her long-term prognosis?* |
| **Q11** | *A patient receiving ipilimumab and nivolumab for metastatic melanoma reports progressive, patchy whitening of his skin without any pruritis or ulceration. Does this impact his prognosis?* |
| **Q12** | *A patient receiving ipilimumab and nivolumab for metastatic melanoma who is responding to treatment develops severe toxicity with hepatitis and colitis and has to stop treatment. How does this impact prognosis?* |
| **Q13** | *A patient with metastatic melanoma who has significant asthenia, weight loss and pain commences combination immunotherapy with nivolumab and ipilimumab. He rapidly improves clinically with reduced pain and increased energy and weight but his first restaging scan reports progression. How should this be managed?* |
| **Q14** | *What germline mutations in cancer are associated with the best prognosis when treated with immunotherapy?* |
| **Q15** | *What is the effect of mismatch repair deficiency in the prognosis and management of endometrial cancer?* |
