## Supplementary material for "Comparison of Large Language Models in Answering Immuno-Oncology Questions: A Cross-Sectional Study": Cohen's Kappa Coefficient for Inter-Rater Reliability

### Supplementary Material C

- Cohen's Kappa Coefficient for Inter-Rater Reliability between the Reviewers during the Selection Process.

| Variable | Cohen’s Kappa | Agreement (%) | Subjects (n°) |
|---|---|---|---|
| Reproducibility | 0.912 | 97.4 | 152 |
| Accuracy | 0.889 | 94.4 | 126 |
| Readability | 1 | 100 | 126 |
| Relevancy | 0.868 | 94.4 | 126 |
