## Supplementary material for "Comparison of Large Language Models in Answering Immuno-Oncology Questions: A Cross-Sectional Study": Numeric data reported in Figure 2 and Figure 3

### Supplementary Material D

- Numeric data reported in Figure 2 (p. 2)
- Numeric data reported in Figure 3. (p. 3)

| **Domain** | **Option** | **GPT-3.5** n (%) | **GPT-4** n (%) | **gBARD** n (%) | GPT-4 vs GPT-3.5 vs gBARD |
|---|---|---|---|---|---|
| Questions with Answers | Overall | 60 (100%) | 60 (100%) | 32 (53.3%) | *p-value = 6.193e-16* |
| | *Mechanisms* | 15 (100%) | 15 (100%) | 14 (93.3%) | |
| | *Indications* | 15 (100%) | 15 (100%) | 5 (33.3%) | |
| | *Toxicities* | 15 (100%) | 15 (100%) | 0 (0%) | |
| | *Prognosis* | 15 (100%) | 15 (100%) | 13 (86.7%) | |

| **Domain** | **Option** | **GPT-3.5** n (%) | **GPT-4** n (%) | **gBARD** n (%) | GPT-4 vs GPT-3.5 vs gBARD |
|---|---|---|---|---|---|
| Reproducibility | Overall | 53 (88.3%) | 57 (95%) | 16 (50%) | *p-value = 8.376e-07* |
| | *Mechanisms* | 13 (86.7%) | 15 (100%) | 6 (40%) | |
| | *Indications* | 13 (86.7%) | 15 (100%) | 1 (6.7%) | |
| | *Toxicities* | 13 (86.7%) | 14 (93.3%) | 0 (0%) | |
| | *Prognosis* | 14 (93.3%) | 13 (86.7%) | 9 (60%) | |

| **Domain** | **Option** | **GPT-3.5** n (%) | **GPT-4** n (%) | **gBARD** n (%) | GPT-4 vs GPT-3.5 vs gBARD |
|---|---|---|---|---|---|
| Accuracy | *Answer rated as "1"* | 4 (7.5%) | 0 (%) | 3 (18.8%) | *p-value = 0.03342* |
| | *Answer rated as "2"* | 18 (34%) | 14 (24.6%) | 6 (37.5%) | |
| | *Answer rated as "3"* | 31 (58.5%) | 43 (75.4%) | 7 (43.8%) | |
| Readability | *Answer rated as "1"* | 0 (0%) | 0 (0%) | 0 (0%) | *p-value = 0.02126* |
| | *Answer rated as "2"* | 1 (1.9%) | 0 (0%) | 2 (12.5%) | |
| | *Answer rated as "3"* | 52 (98.1%) | 57 (100%) | 14 (87.5%) | |
| Relevance | *Answer rated as "1"* | 0 (0%) | 0 (0%) | 1 (6.2%) | *p-value = 0.04107* |
| | *Answer rated as "2"* | 12 (22.6%) | 16 (28.1%) | 8 (50%) | |
| | *Answer rated as "3"* | 41 (77.4%) | 41 (71.9%) | 7 (43.8%) | |
